# Patient-Reported Treatment Outcomes in ME/CFS and Long COVID

**DOI:** 10.1101/2024.11.27.24317656

**Authors:** Martha Eckey, Peng Li, Braxton Morrison, Ronald W Davis, Wenzhong Xiao

## Abstract

Myalgic encephalomyelitis/chronic fatigue syndrome (ME/CFS) and Long COVID are persistent multi-system illnesses affecting many patients. With no known effective FDA-approved treatments for either condition, patient-reported outcomes of treatments are invaluable for guiding management strategies in patient care and generating new avenues for research. Here, we present the results of an ME/CFS and Long COVID treatment survey with responses from 3,925 patients. We assessed the experiences of these patients with more than 150 treatments, as well as their demographics, symptoms, and comorbidities. Patients with each condition who participated in the study shared similar symptom profiles, including all the core symptoms of ME/CFS, e.g., 89.7% of ME/CFS and 79.4% of Long COVID reported post-exertional malaise (PEM). Treatments with the greatest perceived benefits were identified, which had varied effects on different core symptoms. In addition, treatment responses were significantly correlated (R² = 0.68) between the two patient groups. Patient subgroups with distinct profiles of symptoms and comorbidities showed varied responses to treatments, e.g., a POTS-dominant cluster benefiting from autonomic modulators and a cognitive-dysfunction cluster from CNS stimulants. This study underscores the symptomatic and therapeutic similarities between ME/CFS and Long COVID and highlights the commonalities and nuanced complexities of infection-associated chronic diseases and related conditions. Insights from patient-reported experiences, in the absence of approved treatments, provide urgently needed real-world evidence for targeted therapies in patient care and for developing future clinical trials.

(Disclaimer: The findings presented in this paper are based on patient-reported information and are intended for research purposes only. They should not be interpreted as medical advice. Patients are advised to consult their healthcare provider before initiating or altering any treatment.)

## Introduction

Myalgic encephalomyelitis/chronic fatigue syndrome (ME/CFS) and Long COVID are both debilitating, multi-system illnesses that profoundly impact millions of people worldwide (1–5). Long COVID alone currently affects 17.9% of adults in the US (Sept 2024) (6), with many of these patients meeting the diagnostic criteria for ME/CFS (3, 7–9). Before the pandemic, ME/CFS was estimated to affect up to 3.3 million people in the US (1, 10–12), but the surge in Long COVID cases is increasing these numbers significantly. The large socio-economic burden of these conditions will continue to escalate if effective treatments are not identified (12, 13).

Increasing evidence suggests that Long COVID and the majority of ME/CFS cases are infection-associated chronic conditions in which initial infections trigger downstream pathological mechanisms, leading to chronic, multi-system dysfunction (2, 9, 14). Despite the diverse symptoms in individual patients, studies have shown that many Long COVID and ME/CFS sufferers share hallmark symptoms such as fatigue, post-exertional malaise (PEM), unrefreshing sleep, orthostatic intolerance (OI), and cognitive dysfunction. These symptoms align with the Institute of Medicine (IOM) diagnostic criteria for ME/CFS (2, 3, 8, 14–17). Additional symptoms and comorbidities, such as pain (e.g., migraines, neuropathy) and mast cell activation syndrome (MCAS), are also prevalent in both conditions. Furthermore, both conditions occur more frequently in females, with the average age of onset typically in the 30s or 40s (4, 10, 11, 18, 19). These similarities suggest potential links in the underlying pathophysiology of ME/CFS and Long COVID.

Patients with ME/CFS and Long COVID frequently struggle to access specialized medical care (20, 21). Primary care providers and other health professionals often lack the necessary expertise to diagnose and manage ME/CFS or Long COVID effectively, leading to significant delays in appropriate treatments, which negatively impact patients’ physical and emotional well-being. Even when patients do access appropriate medical care, treatment options are limited: With no known cure for Long COVID or ME/CFS, patients must rely on palliative treatments and less accessible off-label medications (22–24). Due to the variety of symptoms and comorbidities found in these conditions, treatments must be uniquely tailored to individual needs, often on a trial-and-error basis. Several systematic reviews summarize the existing clinical evidence of therapies in ME/CFS (25–27). Early reports suggest that many of the treatments—such as low-dose naltrexone (LDN), pyridostigmine (Mestinon), beta-blockers, immunoglobulin therapy (IVIG), and CNS stimulants—may also help improve corresponding symptoms in Long COVID (3, 24, 28, 29). Conversely, Long COVID studies investigating platelet hyperactivation and other prothrombotic changes (30–32) have renewed interest in the use of antiplatelets, anticoagulants, and fibrinolytic supplements in ME/CFS (33–35). As we await further clinical trials, real-world patient reports via surveys can provide quick and direct insights into which therapeutic interventions may or may not be beneficial. Such information has long impacted drug development (36–38) and is particularly important in the management of chronic diseases (39–41). In the absence of approved treatments for ME/CFS and Long COVID, patient-reported treatment outcomes can not only provide urgently needed information for patients and their healthcare providers to help address significant gaps in care but also help identify promising candidates for clinical trials.

Here, we present a comprehensive study of treatment outcomes in 3,925 patients with ME/CFS and Long COVID. The TREATME survey examined over 150 supplements, over-the-counter medications, prescription medications, and non-pharmacological interventions and their perceived effectiveness in managing these debilitating conditions. These treatments were selected based on pharmacological and non-pharmacological therapies currently used in ME/CFS (25–27), various pertinent studies, and conversations with members of the patient community. By assessing the symptoms and comorbidities of patients along with their treatment experiences, we identified therapies with the greatest perceived benefits and the core symptoms they improved. Patient subgroups with distinct symptom profiles also were identified, showing increased responses to specific treatments. This research establishes a foundation for utilizing real-world patient data to better understand therapies for ME/CFS and Long COVID, aiming for targeted treatments that improve patient care.

## Results

### Demographics and Disease Duration of Patients in the Study

3,925 patients responded to the survey and answered questions on demographics, symptom profiles, and comorbidities, including 2,125 patients with ME/CFS and 1,800 patients with Long COVID. As shown in Table 1, 80% of the Long COVID patients (1,440/1,800) and 87.5% of the ME/CFS patients (1,859/2,125) reported that their diagnoses were made formally, while the rest reported that their diagnoses strongly suspected either by the patients themselves or their doctors. Altogether, 76.4% of Long COVID patients and 82.8% of ME/CFS patients were female, and 83.7% of Long COVID patients and 77.2% of ME/CFS patients were between 30 and 65 years old. The average ages of the patients were 47.5 ± 14.9 and 44.1 ± 12.9 for ME/CFS and Long COVID, respectively. These similarities in age and sex breakdown between patient groups roughly align with those reported elsewhere, suggesting that survey respondents are broadly representative of these patient populations.

**Table 1.**
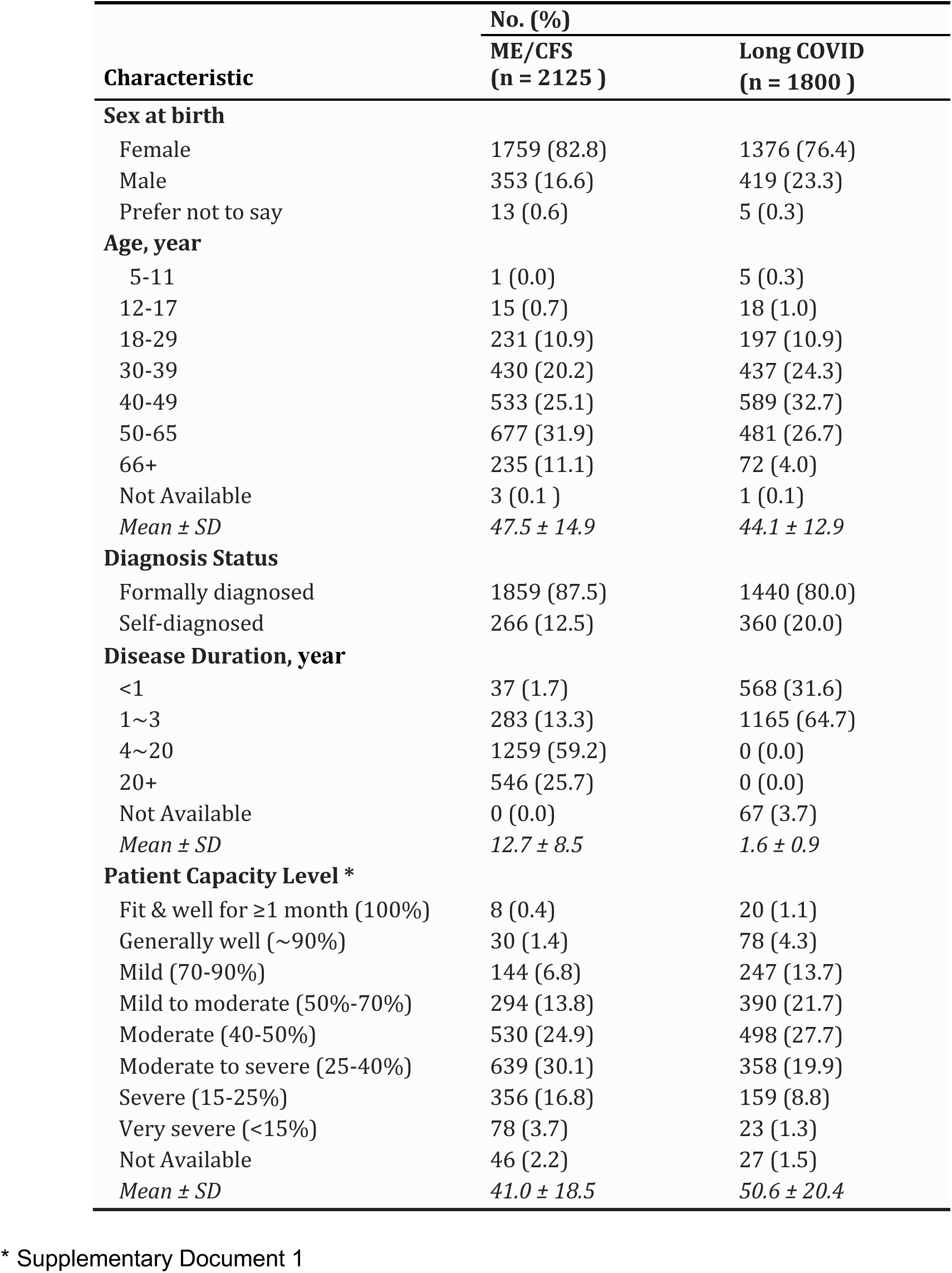
Demographic Characteristics of ME/CFS and Long COVID Patients in the Study.

Given the long history of ME/CFS and the recent emergence of Long COVID, the duration of illness for patients differed widely between the groups. The average disease durations (years) were 12.7 ± 8.5 for ME/CFS and 1.6 ± 0.9 for Long COVID, respectively. While some ME/CFS patients also reported illness durations exceeding 20 years, the length reported by Long COVID patients was naturally capped at 3 years (2020-2023). Both groups reported significantly reduced patient capacity levels (Table 1), and ME/CFS patients had, on average, a lower percentage of their pre-illness capacity compared to Long COVID patients (41.0 ± 18.5% vs 50.6 ± 20.4%). More ME/CFS patients (20.4%) also reported severe (15-25% capacity) to very severe illness (<15% capacity), compared to those with Long COVID (10.1%). Additionally, only 8.6% of ME/CFS patients reported their capacity as ‘mild’ (70-90% capacity)’ or higher, compared to 19.1% of Long COVID patients.

### ME/CFS and Long COVID Patients Share Similar Symptom Profiles

Patients reported the most troubling symptoms experienced over the course of their illness (Supplementary Table 1). Figure 1 shows the top symptoms ranked by their prevalence among respondents with Long COVID. The top five symptoms are fatigue (95.6% of ME/CFS patients, 88.3% of Long COVID patients), post-exertional malaise (89.7%, 79.4%), brain fog (80.1%, 72.3%), unrefreshing sleep (74.5%, 55.3%), and memory problems (54.1%, 50.8%). Both groups also reported high rates of postural orthostatic tachycardia syndrome (POTS; 41.0%, 40.0%), along with other orthostatic intolerance (OI)-related symptoms like fast, fluttering or pounding heartbeat (38.5%, 47.1%), and lightheadedness and dizziness (42.6%, 38.5%). Notably, this symptom profile aligns with the IOM ME/CFS criteria. In addition, both ME/CFS and Long COVID patients had similar rates of insomnia (47.3%, 40.2%), headache or migraine (45.3%, 40.1%), and numbness and tingling (25.4%, 26.4%). In contrast, a lower percentage of ME/CFS patients experienced frequent shortness of breath (dyspnea; 31.5%, 40.4%), chest pain (17.3%, 31.4%), and disordered taste/smell (9.0%, 15.6%), possibly due to the unique respiratory, cardiovascular, and sensory impacts of acute SARS-CoV-2 infection. On the other hand, ME/CFS patients had a higher rate of sore/painful muscles (53.7%, 34.1%).

**Figure 1.**
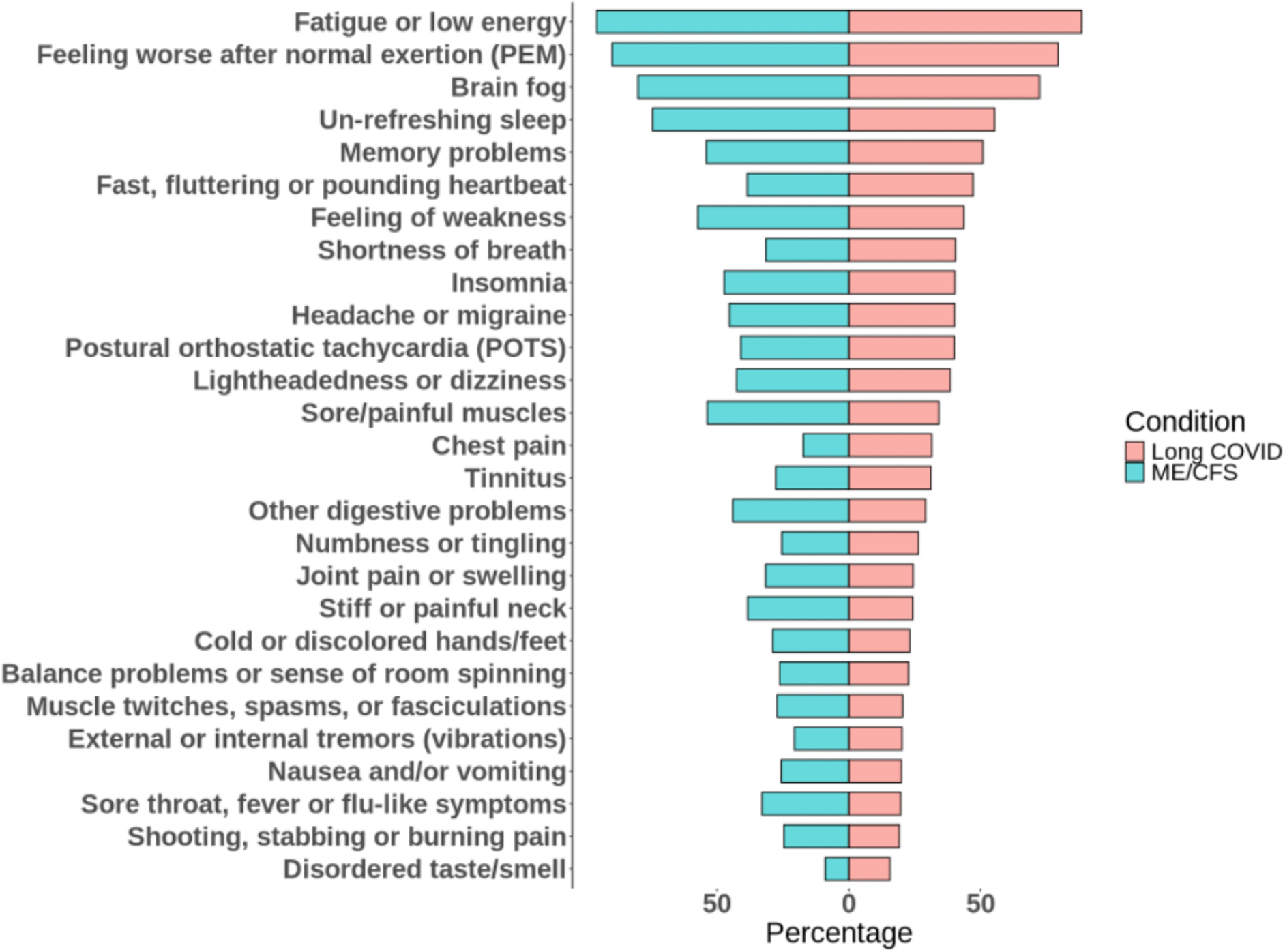
Most Troubling Symptoms in ME/CFS and Long COVID Patients. The bar graph shows the frequency of symptoms reported by patients with ME/CFS (blue) and Long COVID (red), ordered based on the frequency in Long COVID patients.

ME/CFS and Long COVID patients had similar comorbidities, including anxiety and depression (56.6% in ME/CFS, 51.1% in Long COVID), POTS (51.9%, 48.7%), migraine (49.1%, 41.1%), other dysautonomia (37.5%, 38.9%), mast cell activation syndrome (MCAS; 31.8%, 29.1%), Ehlers-Danlos Syndrome (EDS)/joint hypermobility (40.2%, 27.2%), and ADD/ADHD (23.8%, 23.7%) (Supplementary Figure 1). These findings are consistent with other reports suggesting overlapping pathologies of ME/CFS and Long COVID (3, 7, 8, 15, 16, 28, 42, 43), which led us to hypothesize that ME/CFS and Long COVID patients may show similar responses to certain treatments.

### Treatments Identified with the Greatest Perceived Benefit Reported by Patients

The survey included over 440 questions on the use and perceived efficacy of over 150 different interventions. Patients reported how they felt each treatment influenced their overall condition, choosing from seven options: much better, moderately better, slightly better, about the same/unchanged, slightly worse, moderately worse, or much worse. Additionally, patients were asked to choose from those same seven options to evaluate a select treatment’s effects on their most troubling symptoms. The detailed results are shown in Supplementary Data.

Reported effect on overall condition was summarized using a Net Assessment Score (NAS) (see Materials and Methods). Each treatment evaluated by 20 or more patients was assigned a NAS. The NAS of each treatment was then compared to that provided by patients for an oral, non-liposomal Vitamin C supplement—which, in the absence of a placebo, served as a reference—using the false discovery rate adjusted p-value (adj.p) (Figure 2). The results of this analysis for each treatment are in Supplementary Table 2.

**Figure 2.**
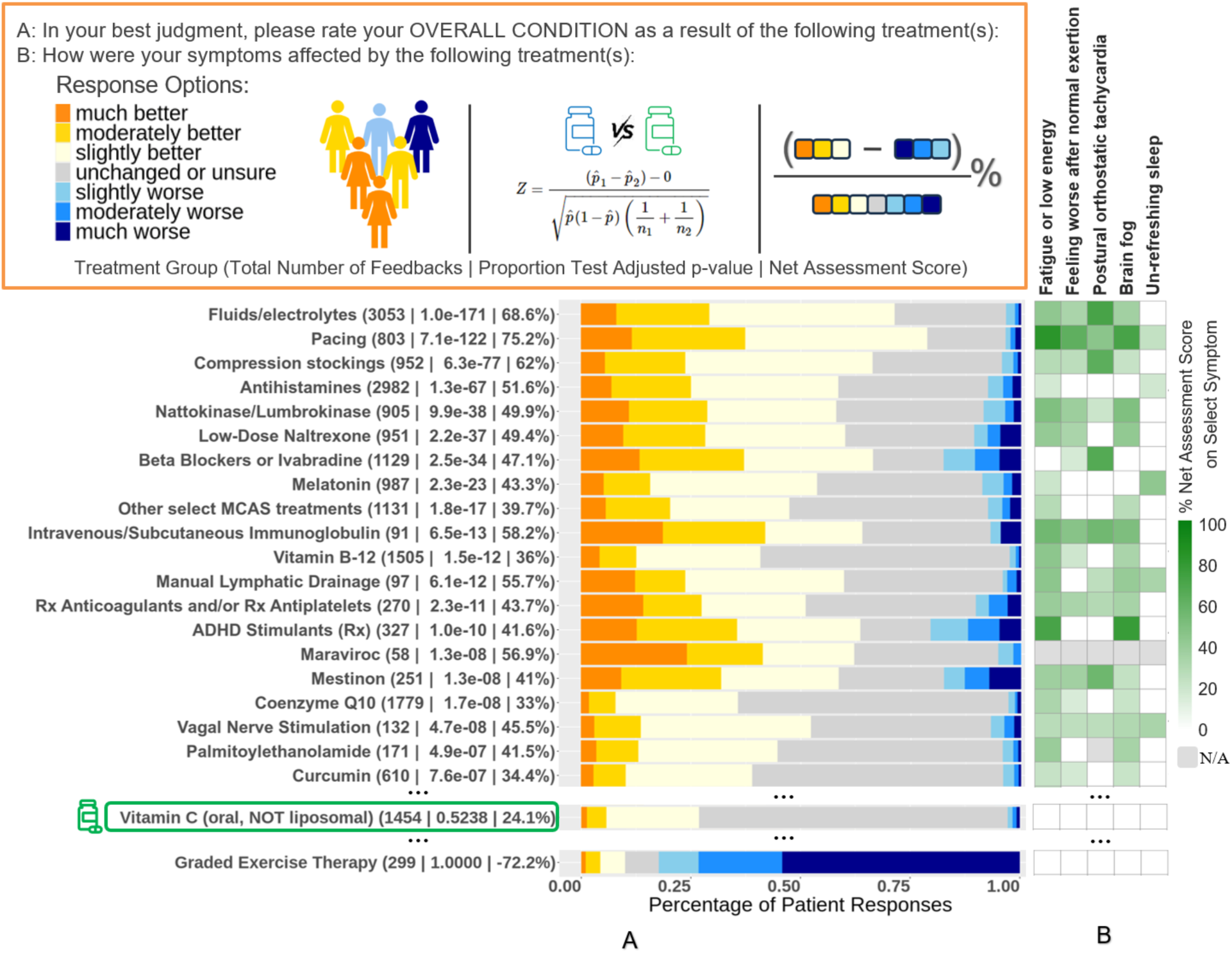
Patient-Reported Outcomes of Treatments in ME/CFS and Long COVID. **A**. The leading treatments and categories that contributed to improving patients’ overall conditions as reported in self-assessments. The data within parentheses next to each treatment indicate the number of responses provided on the treatment, the false discovery rate adjusted p-value compared to the reference treatment, and the Net Assessment Score (NAS). The stacked bar plots show the percentages of patient responses related to the perceived treatment effect of each treatment, spanning from “much worse” to “much better”. Orange shading denotes the percentage of patients who reported a positive treatment effect—with darker orange indicating a stronger positive effect—whereas blue shading indicates adverse side effects, with darker blue representing more significant negative effects. Vitamin C (oral, non-liposomal) was used as the reference treatment. Of all the treatments investigated, Graded exercise therapy (GET) received the lowest NAS score. **B**. Impact of treatments on patient symptoms as reported in self-assessments. For each of the core symptoms, shown in a gradient scale is the proportion of respondents of a treatment who also reported the impact of the treatment improving the specific symptom.

Figure 2A shows the top twenty treatment groups with the greatest perceived benefit reported by patients. For example, fluids/electrolytes demonstrated significantly higher perceived benefit over the reference (adj.p = 1.0e-171) and a NAS of 68.6% based on 3,053 responses. Other treatment groups with the most positive NAS include pacing (75.2%), compression stockings (62%), intravenous/subcutaneous immunoglobulin (IgG; 58.2%), maraviroc (56.9%), manual lymphatic drainage (55.7%), antihistamines (51.6%), nattokinase/lumbrokinase (NK/LK; 49.9%), low-dose naltrexone (LDN; 49.4%), beta blocker or ivabradine (47.1%), vagal nerve stimulation (VNS; 45.5%), Rx anticoagulants/Rx antiplatelets (43.7%), melatonin (43.3%), palmitoylethanolamide (PEA; 41.5%), ADHD stimulants (41.6%), and Mestinon (41%).

Several individual treatments in these treatment groups received high percentages (>40%) of responses indicating “much better” and “moderately better,” including IV saline, ivabradine, IgG, heparin (UFH and LMWH), and maraviroc (Supplementary Table 2). However, a relatively small sample size (<100 patients) evaluated IgG, maraviroc, or heparin. Additionally, B12 injections (NAS of 47.1%) significantly outperformed oral B12 (30.5%), and higher dosages of coenzyme Q10 (50.7% for >200mg/day and 41.0% for 100-200mg/day) were shown to be beneficial while lower dosages weren’t.

It is also important to note that while the majority of patients reported benefits from medications such as beta-blockers, ivabradine, ADHD medications, and Mestinon, a smaller group (17%-21%) indicated that these medications worsened their condition. This highlights patients’ diverse responses to treatments.

Examining other treatments with responses from at least 100 patients (see Supplementary Table 2), corticosteroids (38.4%) and glutathione injection (42.4%) also exhibited significant positive effects. Conversely, a considerable number of other treatments did not show overall significance compared to the reference. Some of these, including Rx antidepressants, gabapentinoids, (ar)modafinil, Abilify, dihydropyridines, fludrocortisone, midodrine, and cognitive behavioral therapy (CBT), had over 20% of patients responding negatively. However, notably, trazodone/nefazodone (antidepressants), pregabalin/gabapentin (gabapentinoids), Abilify ≤ 2 mg (low-dose), and midodrine also had more than 50% of positive responses. This again underscores the diverse responses of patients to treatments. Of all treatments investigated, graded exercise therapy (GET) received the lowest NAS score (−72.2%), with the vast majority of polled patients reporting harms and almost none reporting benefit (Figure 2A).

### Treatments Impact Core Disease Symptoms Differently

As stated earlier, our survey asked patients to evaluate a treatment’s effects not only on their overall condition but also on their most troubling symptoms. The five symptoms we included in our analysis were selected to align with IOM’s core diagnostic criteria for ME/CFS: 1) fatigue or low energy, 2) feeling worse after normal exertion (PEM), 3) POTS, 4) brain fog, and 5) unrefreshing sleep. These symptoms were also among the topmost troubling symptoms selected by survey participants (Supplementary Table 1). Patients were asked to report the extent to which each of their selected symptoms was impacted by each treatment, again selecting from seven options (e.g., “much better”, “slightly worse”, et al.) To investigate the effects of the top 20 treatment groups on these five core symptoms, we calculated symptom-specific NAS scores, which then were compared to the NAS for the reference, vitamin C, to determine which were significantly different. Figure 2B shows the frequencies of a treatment that significantly improved each symptom compared to vitamin C (adj. p-value < 0.05).

Each treatment affects the symptoms differently. For example, patients reported that pacing helped frequently with fatigue (82.7%), PEM (62.6%), brain fog (71.2%), and POTS (45.5%). Similarly, IgG improved these symptoms frequently: fatigue (55.4%), PEM (46.7%), POTS (55.8%), and brain fog (50.8%). Fluids/electrolytes most frequently helped POTS (71.7%), and to a lesser extent, fatigue or low energy (43.2%), PEM (33.2%), and brain fog (36.5%). As expected, compression stockings and Mestinon most frequently improved symptoms of POTS (63.7% and 56.9%, respectively), while beta blockers and ivabradine primarily alleviated POTS symptoms (66.0%). LDN, on the other hand, did not improve POTS but significantly improved three other core symptoms: fatigue or low energy (41.5%), PEM (33.2%), and brain fog (42.3%). Interestingly, NK/LK similarly improved these three symptoms (49.7%, 38.2%, and 49.5%, respectively), and also showed some benefits in POTS (20.3%). As expected, ADHD stimulants significantly alleviated brain fog (77.1%) and general fatigue (71.7%) but did not improve PEM. Among the treatments surveyed, only melatonin significantly improved unrefreshing sleep (43.0%).

These results indicate that treatments have varying effects on disease symptoms, suggesting that patients potentially can benefit from different treatment options based on their individual needs. This is further supported by the observation that patients with specific comorbidities (e.g., OI) have used particular treatments that contributed to improving their symptoms (e.g., Mestinon) (Supplementary Figure 2).

### Responses to Treatments are Similar between ME/CFS and Long COVID

To compare the treatment response between the two conditions, we performed analyses of ME/CFS and Long COVID patients separately (see Materials and Methods). As shown in Figure 3, the NAS scores of treatments were highly correlated between ME/CFS and Long COVID patients (R² = 0.68), indicating similarities in treatment responses. This correlation was particularly pronounced for treatments reported by larger numbers of ME/CFS and Long COVID patients. For complete results, see Supplementary Table 3. However, overall, Long COVID patients often reported more positive response, i.e., higher NAS, to many treatments. Still, only two treatment groups—midodrine, benfotiamine or thiamine tetrahydrofurfuryl disulfide (TTFD)—showed significantly different responses between the two conditions (adj. p <0.05 and fold change > 1.25).

**Figure 3.**
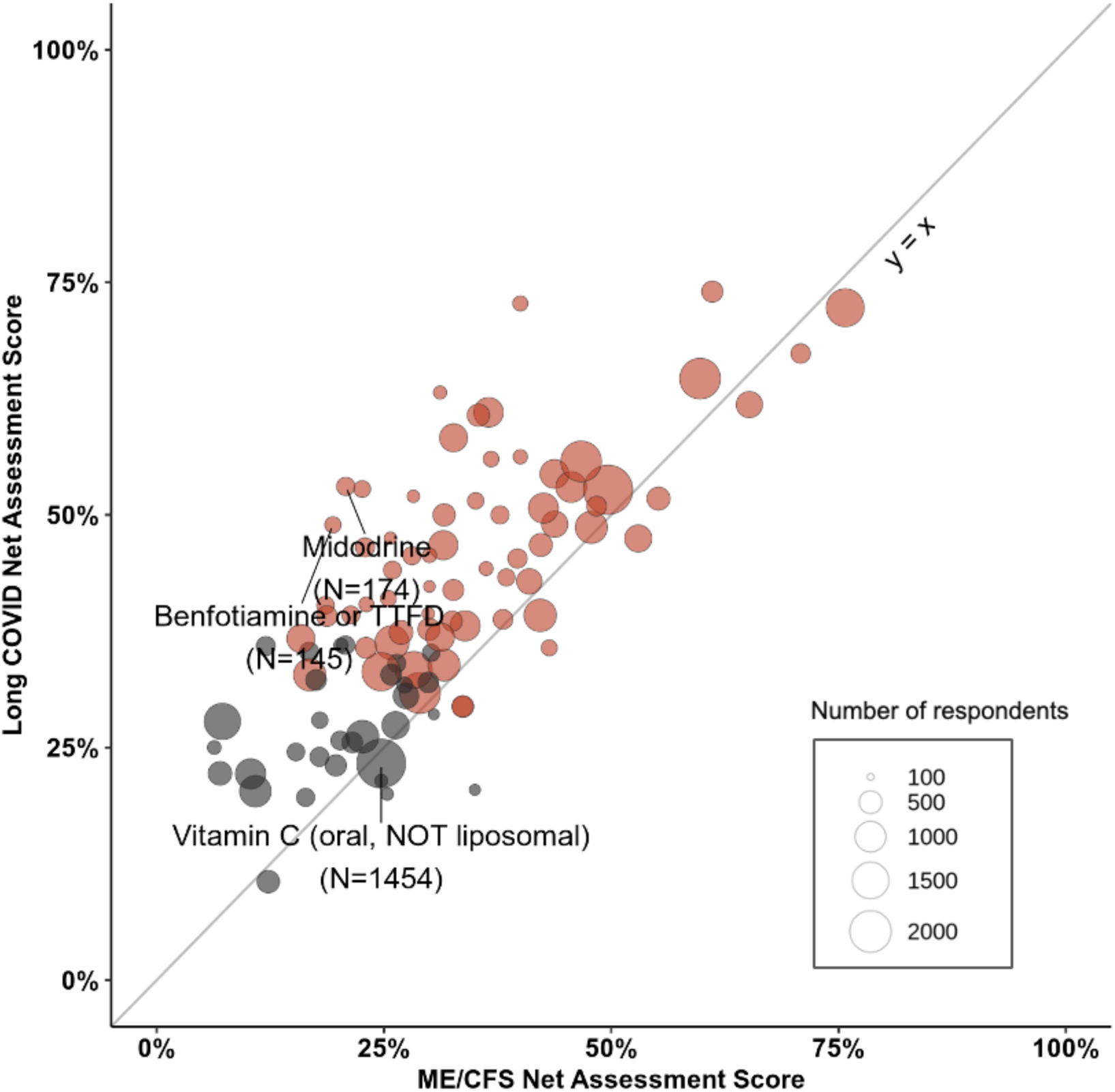
Comparison of Treatment Effectiveness Reported in ME/CFS and Long COVID. Shown is the scatter plot of the Net Assessment Scores (NAS) of treatments in ME/CFS (x-axis) compared to Long COVID (y-axis), with each circle symbolizing a treatment. Red circles represent treatments that showed significance when compared to the reference (Vitamin C; oral, NOT liposomal), and gray circles indicate otherwise. The size of a circle represents the total number of respondents to the treatment.

To further explore the differences seen between ME/CFS and Long COVID patients, we identified significant predictors of treatment effectiveness, i.e., the NAS score, from disease diagnosis (ME/CFS vs. Long COVID), severity (patient capacity level), and demographics of patients (Supplementary Figure 3). Disease severity, i.e., patient capacity level, was by far the most significant predictor of treatment effectiveness. Of the remaining variables, sex, disease duration, diagnosis status, and age all show a greater influence on the treatment outcome than did the patient’s diagnosis of ME/CFS or Long COVID. Therefore, the diagnosis of Long COVID and ME/CFS alone does not impact responses to treatments significantly.

### Subgroups of Patients Had Distinct Profiles of Symptoms and Comorbidities

Since the survey results show that treatments impact disease symptoms differently, we further clustered the symptoms and comorbidities of the survey respondents (see Materials and Methods). Patients formed four clusters with distinct profiles, as further discussed below (Figure 4A).

**Figure 4.**
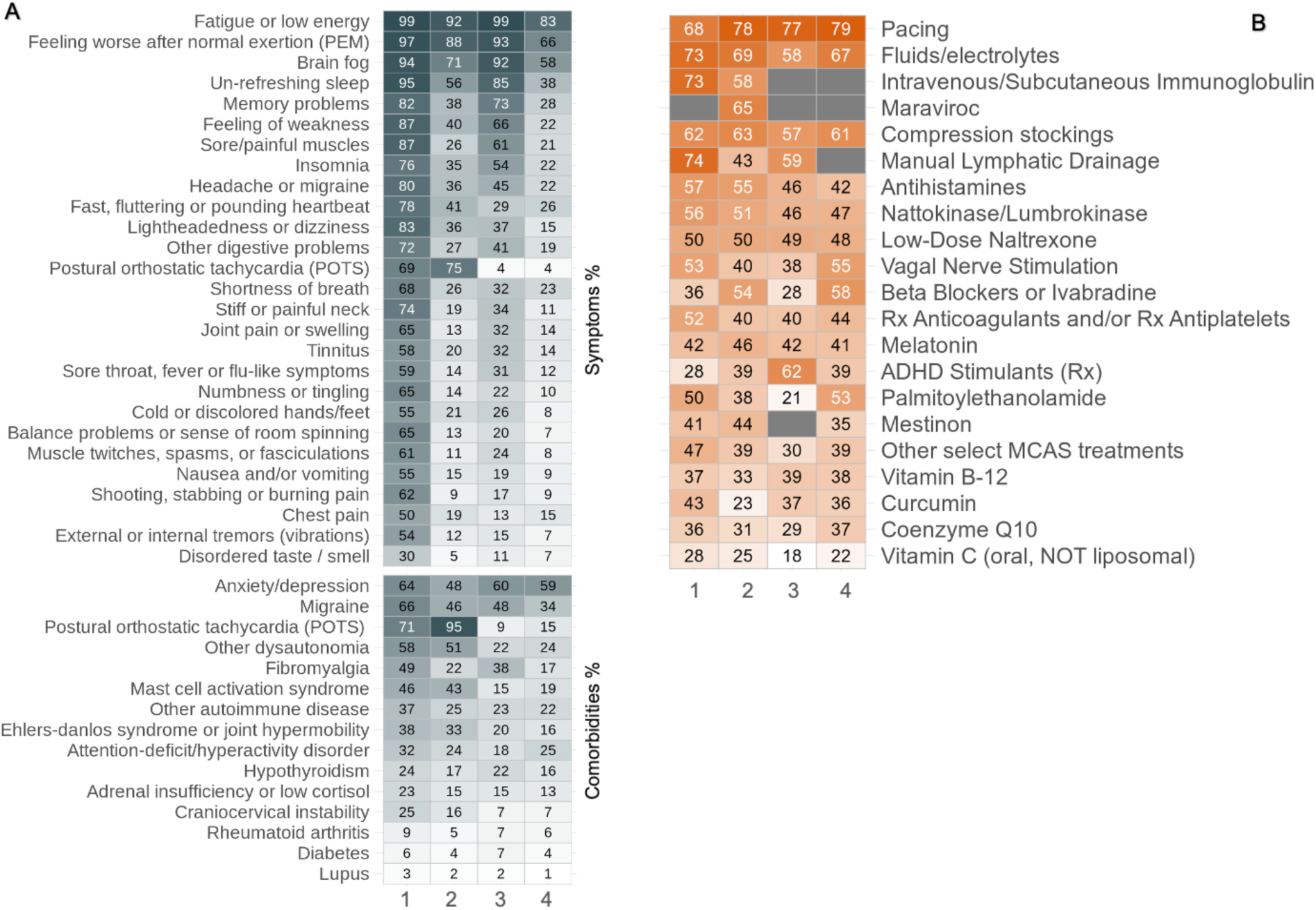
Patients Subgroups with Distinct Symptom Profiles and Treatment Efficacies. **A.** The heatmap displays the percentages of reported symptoms and comorbidities within each patient cluster. Cluster 1 had the highest prevalence of symptoms and comorbidities, Cluster 2 was predominated by postural orthostatic tachycardia syndrome (POTS) issues, Cluster 3 was characterized by cognitive symptoms like brain fog and sleep disturbances, and an increased reporting of pain, and Cluster 4 had the mildest symptoms. **B.** The heatmap shows the comparative effectiveness of treatments across the four distinct patient clusters. The treatments listed are consistent with those reported in Figure 2, and the NAS for each treatment in patients of each cluster are shown. The colors indicate treatment efficacies within each cluster, with warmer colors (reds and oranges) representing higher positive impact, and cooler colors (blues) representing lower positive impact. Grey denotes insufficient feedback (<20) for reliable NAS calculation.

Cluster 1: Multisystemic Symptomatology. Cluster 1 was characterized by higher rates of nearly all symptoms and comorbidities than those reported for every other cluster. Patients in this cluster also reported the lowest functional capacity level (39.08% +/- 17.63, Supplementary Figure 4). As expected, the most prevalent symptoms are the core symptoms of the disorder: fatigue (99.0%), PEM (97.4%), brain fog (93.6%), unrefreshing sleep (95.2%) sore/painful muscles (87.0%), and headache/migraine (79.8%). Patients in cluster 1 also showed higher rates of almost every queried symptom and comorbidity than other clusters. The exceptions to this trend are POTS symptoms (69.0% cluster 1; 75.0% cluster 2), POTS diagnosis (71.0% cluster 1; 95.1% cluster 2), and diabetes (5.96% cluster 1; 6.63% cluster 3).

Cluster 2: POTS-Dominant Presentation. Cluster 2 was primarily marked by the highest rates of POTS reported as a primary symptom (75.0%) and comorbidity (95.1%). These patients also report lower prevalences of most primary symptoms compared to Cluster 3, except fast, fluttering, or pounding heartbeat (77.9%) and chest pain (18.7%). However, compared to Cluster 3, patients in Cluster 2 reported higher rates of certain conditions, including dysautonomia (50.5%), MCAS (42.6%), EDS (33.5%), and craniocervical instability (16.0%). The rates of adrenal insufficiency or low cortisol, lupus, and other autoimmune diseases between these clusters were similar.

Cluster 3: Cognitive and Sleep Dysfunction with Increased Pain. Patients in Cluster 3 reported far higher levels of fatigue, PEM, brain fog, and unrefreshing sleep than those in Clusters 2 and 4, with similarly high levels of fatigue, PEM, and brain fog to those reported by patients in Cluster 1. However, they reported markedly low rates of POTS symptoms (3.73%), similar to those in Cluster 4 (3.97%). Notably, the comorbidities for which Cluster 3 patients reported higher levels than those in Cluster 2 are predominated by pain conditions, including migraine (45.6% Cluster 2; 48.5% Cluster 3), fibromyalgia (22.2% Cluster 2, 37.9% Cluster 3), and rheumatoid arthritis (4.6% Cluster 2, 7.2% Cluster 3). As expected, these patients also reported higher levels of pain symptoms than those in Cluster 2.

Cluster 4: Milder Symptomatology. Cluster 4 included patients with the mildest symptoms and highest mean capacity scores (57.50 +/- 20.6), indicating relatively preserved functionality. Compared to other clusters, members of this cluster have lower average severity of fatigue, PEM, brain fog, and unrefreshing sleep compared to the other clusters, as well as markedly low rates of POTS symptoms.

Separating respondents into those with ME/CFS and Long COVID had minimal effect on the distribution of symptoms within each of the four clusters (Supplementary Figure 5).

### Perceived Effectiveness of Specific Treatments Differs between Patient Subgroups

After defining separate symptom clusters, we next investigated whether survey respondents in the different clusters reported different responses to particular treatments. Figure 4B shows that the treatments identified as having the greatest perceived benefits above—pacing and fluids/electrolytes—are effective across all the patient clusters. This is also consistent with the broad effect of these treatments on multiple core symptoms, as described above (Figure 2B).

However, other treatments showed varying effects across different clusters of patients. In Cluster 1, which consisted of patients with the most symptoms and comorbidities, IgG (73%) and manual lymphatic drainage (74%) had the highest positive response rates, together with Fluids/Electrolytes (73%) and pacing (68%). In Cluster 2, where patients had POTS-dominant presentation, pacing (78%), fluids/electrolytes (69%), maraviroc (65%), and compression stockings (63%) received the highest NAS scores. For patients in Cluster 3, who experienced cognitive and sleep dysfunction and increased pain, ADHD Rx medications (62%) were reported as beneficial, in addition to pacing (77%). However, patients in Cluster 1 reported no significant improvement from ADHD Rx medications to the reference. In Cluster 4, patients who had milder symptoms reported pacing (79%) and fluids/electrolytes (67%) as the most effective.

These findings suggest that understanding a patient’s specific symptom profile may enable clinicians to tailor more effective therapeutic strategies.

## Discussion

Our study uncovers striking similarities in the demographics, symptom presentation, comorbidities, and treatment outcomes between Long COVID and ME/CFS patients. These findings highlight opportunities for integrative research on disease mechanisms and clinical care for both conditions (2).

Long COVID and ME/CFS patients in the study showed similar age and sex distribution, in addition to sharing most of the same symptoms and comorbidities, with few exceptions (Figure 1). Both groups reported the same top four most frequent symptoms: fatigue, PEM, brain fog, and unrefreshing sleep, as well as high rates of OI-related symptoms, which match IOM ME/CFS diagnostic criteria(1). Other symptoms—memory problems, insomnia, dyspnea, headache, sore/painful muscles, numbness, and tingling—and comorbidities like anxiety and depression, POTS, migraine, other dysautonomia, EDS/joint hypermobility, MCAS, and ADD/ADHD, were also commonly present in both conditions. Importantly, responses to the broad range of treatments were also highly correlated between ME/CFS and Long COVID patients (Figure 3). Taken together, these findings are part of mounting evidence indicating overlapping pathologies of these illnesses (3, 7–9, 15).

Given that many ME/CFS patients have struggled with the illness for decades, it is extremely concerning that many of the patients with Long COVID in the survey met the diagnostic criteria for ME/CFS, as it suggests Long COVID may pose a significant, long-term public health burden unless better treatments are developed for these disease conditions. This circumstance is particularly critical as early diagnosis and treatment for ME/CFS—and likely for Long COVID—are critical to slowing or preventing the worsening of symptoms and deterioration of patient health (44).

Because of the lack of approved treatments for ME/CFS and Long COVID, there is an urgent need for information that can help patients and their healthcare providers manage these debilitating conditions and identify potential candidates for clinical trials. In this study, we gathered and assessed patient-reported outcomes from a wide variety of over 150 therapeutic interventions. To reduce potential biases, we chose one of the commonly used treatments, a Vitamin C supplement (oral, non-liposomal), as a reference, and the outcome of each treatment perceived by patients was compared with that of this reference.

Many of the treatments among the top 20 (Figure 2A) are supported by consensus guidelines (22–24), indicating this approach may serve as a novel method for assessing real-world evidence of therapies and identifying promising candidates for clinical trials. For example, pacing is recommended in ME/CFS patient care to manage energy levels and avoid PEM; fluid/electrolytes, compression stockings and Mestinon to help manage OI; beta-blockers and ivabradine to aid tachycardia present in POTS; IgG is used in immune dysfunction; antihistamines (H1 and/or H2 inhibitors) and other select treatments in MCAS; low-dose naltrexone treats pain; and melatonin for sleep problems. Our results also showed that each of these treatments had varying effects on different symptoms (Figure 2B), underscoring the importance of managing these conditions based on patients’ symptoms and needs.

Other treatments in the top 20 provide evidence for considering these therapies when managing patient symptoms. For example, vitamin B12 deficiency is known to cause fatigue and other ME/CFS-like symptoms(22, 44), and B12 injections significantly outperformed oral B12 in helping patients, consistent with a previous study(45). Similarly, higher dosages of coenzyme Q10 were also found to be significant, consistent with a recent trial in Long COVID(46). Both treatments improved fatigue, PEM, and brain fog, but not POTS (Figure 2B). PEA, which has been investigated for pain control and reducing neuroinflammation (47), improved fatigue and brain fog. In addition, vagal nerve stimulation and manual lymphatic drainage are non-pharmacological approaches that helped patients with either condition.

Thrombotic sequelae have been investigated in Long COVID (3, 30–32). In this study, nattokinase/lumbrokinase and Rx anticoagulants/Rx antiplatelets were both shown to significantly help with multiple symptoms, except sleep problems, in patients with Long COVID and ME/CFS. Anxiety/depression and ADD/ADHD were notable comorbidities associated with both conditions in the study, and ADHD stimulants (Rx, e.g., Vyvanse and Adderall) were found to significantly improve brain fog and fatigue but not PEM or POTS. Maraviroc, a CCR5 co-receptor antagonist, also had a high percentage of positive responses (from a limited group of 58 patients, mostly with Long COVID). On the other hand, a considerable number of treatments evaluated did not have patient-reported outcomes significantly better than the reference, partially due to negative responses from a portion (>20%) of patients for some of these treatments. Further studies are required to investigate these findings and develop and adjust treatment strategies accordingly.

Since each treatment alleviates particular symptoms, identifying specific patient subsets is crucial for effective treatment. Four clusters of patients were identified in this study, characterized by 1. multisystemic symptomatology, 2. POTS-dominant presentation, 3. cognitive and sleep dysfunction with increased pain, and 4. milder symptomatology (Figure 4A). While pacing and fluids/electrolytes produced improvement across all clusters, specific treatments had the highest positive response rate in particular clusters (Figure 4B). For example, ADHD Rx medications were most effective in Cluster 3, while they were no better than the reference in Cluster 1. Our findings represent an early effort to identify traits of patients most likely to benefit or be harmed by medications, which can eventually assist in establishing effective criteria or algorithms to determine which patients may benefit in clinical settings, i.e., precision medicine.

Clinical trials in ME/CFS and Long COVID are urgently needed. Therapeutic candidates for randomized controlled trials (RCTs) for ME/CFS are typically identified through a combination of anecdotal patient self-reports, clinical observations, chart reviews, case reports, and open-label clinical trials (OLCTs)(48–52). This study represents the first iteration of a new approach that utilizes patient-driven surveys to assess outcomes of a large number of treatments, aiming to identify promising candidates for RCTs and specific patient subsets that may benefit from these treatments. Online treatment surveys can reach a wider, more diverse patient population than is typically feasible with other approaches. For example, case reports and OLCTs are typically limited to single or few specialty clinics where patients tend to share referral-driven characteristics and are well enough to reach the clinic. Because of the absence of objective biomarkers for Long COVID or ME/CFS, most treatment studies to date are restricted to patient-reported outcomes. Large-scale survey studies that collect patient-reported treatment outcomes together with patients’ demographic and clinical characteristics—such as comorbidities, clinical signs, symptoms, and disease severity—are particularly effective at determining whether certain treatments work for specific subsets of patients. The findings from this study support this approach. Considering the significant diversity among patients with these complex disease conditions, the development of clinical trials focused on targeted treatments may prove to be the most impactful strategy overall.

Our study has several important limitations. First, due to the lack of randomization, the presence of confounders can affect the results of our study, similar to those found in case reports and OLCTs. For example, treatment outcomes may be influenced by the placebo effect, which can vary based on the expectation of outcomes from specific treatments, and some patients may experience spontaneous improvement. Further, patients might trial multiple treatments at once, making it difficult to attribute changes in patient conditions to a particular treatment. In this study, we attempted to offset some of these influences using Vitamin C (oral, non-liposomal) as a reference group. Second, while an online patient survey can reach a larger number of patients from a more diverse patient population, data collected are likely more prone to noise compared to case reports or OLCTs, where health professionals or researchers record treatment details. Future studies can benefit from integrating electronic health records (EHR) to confirm the clinical characteristics and treatments of participating patients. Third, although the current study included nearly 4,000 patients, many treatments still have fewer than 100 responses, which limits the reliability of the estimates of treatment outcomes. Further targeted surveys are needed to confirm these results. By utilizing prospective surveys, health-tracking apps, EHR integration, and increased patient education, future studies will allow us to extend our findings while increasing and verifying the accuracy of collected data.

In conclusion, this research establishes a foundation for utilizing patient-reported experiences from a large-scale treatment survey to provide real-world evidence for therapies in patient care and to inform candidates for future clinical trials in ME/CFS and long COVID.

## Materials and Methods

### Design of the TREATME Survey

The TREATME questionnaire was designed to survey individual experiences with a diverse spectrum of treatments used by ME/CFS and Long COVID patients. The survey is available online (at https://www.surveymonkey.com/r/treatme_v2). Briefly, the first section of the survey includes questions on patient demographics, diagnoses, illness duration and severity, symptoms, comorbidities, and lab tests, while the remaining sections focus on treatments. Surveyed treatments were selected based on pharmacological and nonpharmacological therapies used in ME/CFS (25–27), emerging and pertinent clinical studies of Long COVID, ME/CFS, and related comorbidities, and extensive discussions with patient community members. For example, antiplatelets, anticoagulants, and fibrinolytic supplements (e.g., nattokinase or lumbrokinase) were surveyed in part based on data suggesting platelet hyperactivation and other prothrombotic changes in Long COVID patients (30–32), and other literature suggests the presence of coagulopathies in ME/CFS as well (33–35). Guanfacine, in combination with N-acetylcysteine, was surveyed based on neurobiological data and case reports showing cognitive benefits in Long COVID (53). Orally administered non-liposomal vitamin C served as a comparator agent against which all other treatments could be measured. The use of oral vitamin C in this way was motivated by its widespread use, limited oral absorption (54), and inability to reach anywhere near the supratherapeutic concentrations which have shown promise in EBV “CFS” patients(55) or acute COVID inpatients (56).

In total, the survey included over 440 questions on the use and perceived efficacy of over 150 medications, supplements, and other interventions organized into 25 broader treatment categories. To borrow strength from treatments with similar modes of action, these treatments were consolidated into 97 treatment groups for downstream analysis. To encourage more accurate results, patients were asked to review treatments only if they had a good idea of their effects, whether positive, negative, or neutral. Patients reported how they felt each treatment influenced their overall condition, choosing from seven options: much better, moderately better, slightly better, about the same/unchanged, slightly worse, moderately worse, or much worse. Additionally, patients were asked to choose from those same seven options to evaluate a select treatment’s effects on their most troubling symptoms; i.e., symptoms individually selected near the survey’s beginning and later piped into specific treatment questions. Patients were also asked about side effects, duration of treatment, length of time before perceived benefit (if any), and duration of benefit (if present). When particularly pertinent, various questions about dosage, specific formulations, and supplement brands also were included. Patients were able to pause and resume the survey throughout the process and as many times as needed; the progress on the survey was stored in each respondent’s web browser.

### Contributions from the Patient Community in the Development of the Survey

Due to difficulties accessing appropriate medical care or effective treatments, many Long COVID and ME/CFS patients suffering from these conditions have turned to social media to exchange information, discuss treatments, and even run their own patient-led experiments. Our survey grew out of discussions with a large cross-section of these individuals. Over a year prior to the creation of this survey, its author began releasing smaller surveys and polls on Twitter, gathering information on the efficacy of popular or controversial treatments used in the patient community. As the surveys gained traction, an increasing number of respondents provided suggestions and feedback which ultimately were utilized in the development of this survey.

Before the survey was released publicly, over 20 individuals with Long COVID and/or ME/CFS took the survey as a trial run so that further feedback could be collected. Some expressed worries about the survey’s length causing fatigue or post-exertional malaise, so optional breaks were included throughout the survey, and both skip logic and advanced piping were employed to limit question burden. Trial respondents also stressed the importance of including “slightly” better/worse response options (rather than only “moderately” and “much” better/worse, which also had been considered), because even a “slight” improvement in symptoms from a given treatment made a significant impact on their quality of life, and oftentimes, improvements greater than “slight” were never reached. Thus, including the response option of “slightly” better/worse ensured a greater breadth of information was collected. Furthermore, it was also noted that sometimes treatments would work well for a while, but then inexplicably stop working, so a recurring question was added to clarify if benefits were sustained or temporary. Such feedback from the community proved invaluable for survey construction.

### Data Collection of the TREATME Survey

Data were collected through an online survey (on surverymonkey.com), which was distributed to patients with either condition, from February 2023 to July 2023 (round 1) and October 2023 to February 2024 (round 2). 5,451 responses were received. Responses without a confirming answer on either ME/CFS or Long COVID diagnosis (130) and/or those without answering any questions on treatments (1,295) were excluded from further analysis. For participants who responded in both rounds of the survey (101), only their first-round responses were retained for analysis. These resulted in 3,925 responses for downstream analysis, comprising 2,125 patients with ME/CFS and 1,800 with Long COVID.

### Evaluation of the Effect of Treatments from Patients’ Responses

To evaluate patient-reported effects of treatments, we defined a Net Assessment Score (NAS) based on patients’ responses on the impact of each treatment on their overall condition:

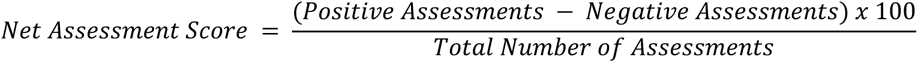

The number of positive and negative feedback correspond to patients reporting any level of improvement or worsening, respectively. The total number of respondents reflects those who have tried the treatment and contributed feedback.

We also calculated the NAS score of a treatment specific to the impact on each core symptom: fatigue or low energy, feeling worse after normal exertion (PEM), POTS, brain fog, and unrefreshing sleep.

For each treatment with a sample size of at least 20, the NAS for the overall condition or a specific symptom was then compared to that provided by patients for a Vitamin C supplement (oral, non-liposomal) as a reference.

The difference between the treatment and the reference group (vitamin C;oral, NOT liposomal) was analyzed using a two proportion Z test(57), which compares the proportion of improvement reported in each group. The test was applied as follows:

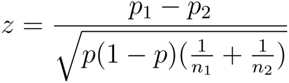

Where p1 and p2 represent the proportions of reported improvement (NAS) in the treatment and vitamin C reference groups, respectively, p is the combined proportion of improvement, and n1 and n2 are the sample sizes. The test statistic z follows a standard normal distribution, and the one-sided p-value indicates the probability of observing a proportion of improvement at least as extreme as p1 under the null hypothesis that the treatment is not better than the vitamin C reference group. Adjusted p-values were controlled using the Benjamini-Hochberg procedure to adjust for multiple tests across various symptom reports, and a significant effect is defined as adjusted p-value (adj. p) <0.05.

### Comparison of Treatment Response Between ME/CFS and Long COVID

To identify treatments with significantly different effects between ME/CFS and Long COVID patients, a two-proportion Z-test was performed on the NAS of the treatment in the two conditions and the fold change between the scores. Only treatments taken by both ME/CFS and Long COVID patients with a total sample size of over 100 were included in this analysis to reduce the impact of random effects on treatments with limited feedback. Treatments meeting abs(fold change) > 1.25 and multiple tests adjusted p-value <0.05 were identified as significant between the two conditions.

To compare the impact of different factors on reported treatment efficacy, significant predictors of treatment effectiveness (i.e., the NAS) were identified from disease diagnosis (ME/CFS vs Long COVID), severity (patient capacity level), and demographics of patients. A gradient boosting machine (GBM) model (58) was employed, utilizing a Gaussian loss function. It included 5000 trees and was validated using 5-fold cross-validation.

### Subclustering of Patients Based on Symptoms and Comorbidities

Uniform manifold approximation and projection (UMAP) (59, 60) was used to transform the high-dimensional patient data, encompassing demographic information, symptoms, and comorbidities into a two-dimensional space. K-means clustering was then applied to cluster patients into subgroups. The optimal number of clusters was determined as 4 (Supplementary Figure 6). For each treatment with a sample size greater than 20 in a cluster, the NAS for that treatment was calculated within the cluster.

## Data Availability

All data produced in the present work are contained in the manuscript

## Acknowledgments

The survey grew out of discussions with many patients in the ME/CFS and Long COVID communities, and numerous patients provided invaluable feedback throughout the project. We would also like to express our sincere gratitude to the more than 5,000 patients who participated in this study. This study would not have succeeded without the tremendous support from the patient communities. We would also like to extend a special thank you to Dr. Trent Garrison, who helped Dr. Eckey with critical steps behind the scenes. Additionally, we appreciate the helpful advice we received from physicians specializing in ME/CFS and Long COVID.

This work was partially supported by Open Medicine Foundation (WX).

## Author Contributions

ME and WX conceived and designed the study. WX and RWD supervised the project. ME designed the survey and collected the dataset, ME, PL, BM, and WX revised the survey. PL, ME, BM performed the data analysis and conducted the statistical analysis. PL, ME, BM, WX and RWD contributed to the interpretation of the results. BM, PL, ME and WX drafted the manuscript. ME and RWD revised the manuscript. All authors read and approved the final manuscript.

