## Supplementary Figures for "Patient-Reported Treatment Outcomes in ME/CFS and Long COVID"

**Supplementary Figure 1: Rates of Comorbidities in ME/CFS and Long COVID Patients**

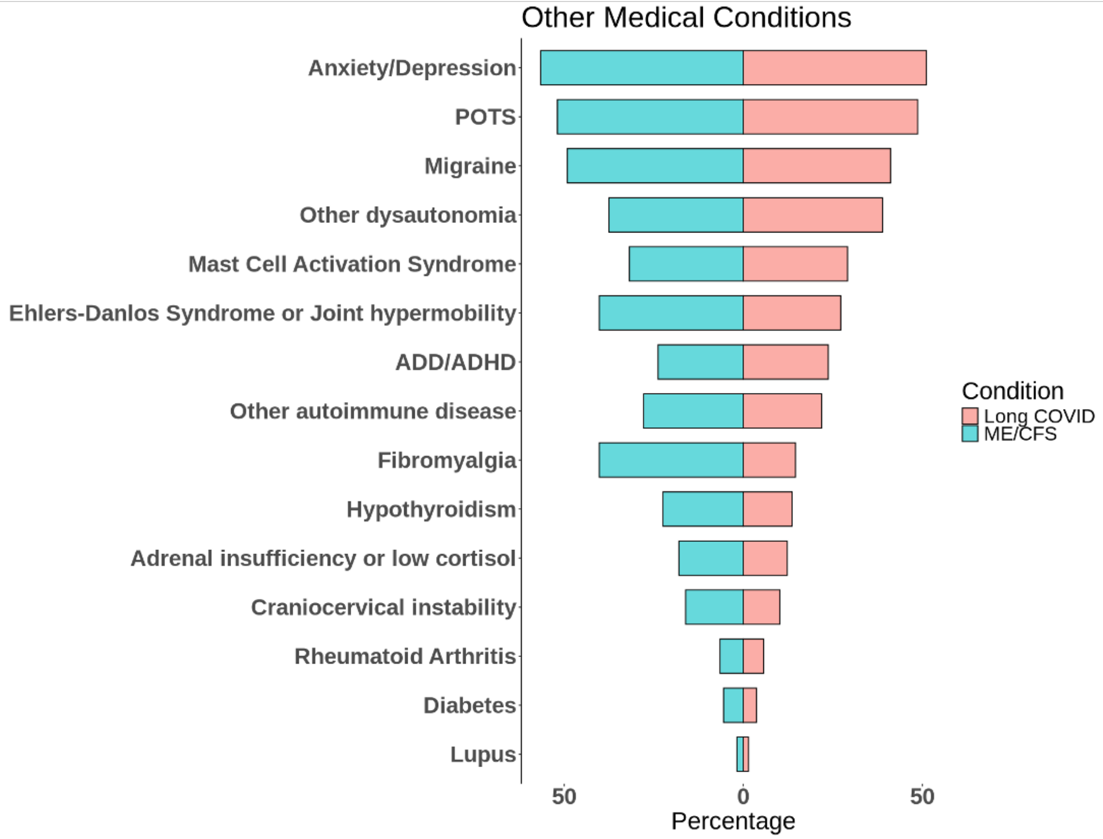

**Supplementary Figure 2**: **Rates of Comorbidities among Patients Reported Using Each Treatment**

**Heatmap of Treatment Group Percentages**

| Treatment | Factor V Leiden | Heart failure | Lupus | Heart inflammation (example: myocarditis) | Antiphospholipid Syndrome | Kidney disease or damage | Clotting disorder | Liver disease or damage | Diabetes | Lung disease or damage | Rheumatoid Arthritis | Preload failure | Other clotting disorder | Immune dysfunction | High blood pressure |
|---|---|---|---|---|---|---|---|---|---|---|---|---|---|---|---|
| Fluids/electrolytes | 0.6 | 0.8 | 1.5 | 1.0 | 1.4 | 1.2 | 1.2 | 2.5 | 3.9 | 3.6 | 4.4 | 4.7 | 6.4 | 8.6 | 11.0 |
| Pacing | 0.0 | 3.0 | 0.9 | 3.7 | 0.0 | 4.4 | 4.6 | 6.8 | 5.4 | 11.5 | 4.1 | 0.0 | 0.0 | 26.9 | 0.0 |
| Compression stockings | 0.4 | 0.9 | 1.8 | 1.1 | 1.7 | 0.9 | 1.3 | 2.7 | 2.8 | 3.6 | 4.8 | 5.0 | 7.1 | 7.9 | 12.3 |
| Antihistamines | 0.6 | 0.6 | 1.6 | 0.8 | 1.8 | 1.1 | 1.4 | 2.1 | 3.3 | 2.9 | 5.2 | 4.8 | 7.7 | 7.5 | 15.0 |
| Nattokinase/Lumbrokinase | 1.1 | 0.0 | 1.6 | 0.6 | 2.0 | 0.1 | 0.8 | 0.6 | 1.7 | 0.7 | 4.8 | 4.8 | 12.1 | 1.4 | 17.1 |
| Low-Dose Naltrexone | 0.8 | 0.9 | 1.3 | 1.5 | 2.2 | 2.0 | 2.3 | 3.2 | 4.3 | 4.0 | 5.8 | 5.0 | 5.3 | 11.9 | 11.7 |
| Beta Blockers or Ivabradine | 0.6 | 0.7 | 2.1 | 1.2 | 1.5 | 1.5 | 1.5 | 2.5 | 4.2 | 3.7 | 5.3 | 4.9 | 6.0 | 8.4 | 15.8 |
| Melatonin | 1.0 | 0.0 | 1.9 | 0.0 | 2.7 | 0.0 | 0.0 | 0.0 | 3.8 | 0.0 | 6.9 | 6.4 | 10.5 | 0.0 | 20.5 |
| Other select MCAS treatments | 1.2 | 1.0 | 1.7 | 1.0 | 2.4 | 1.2 | 1.7 | 3.0 | 3.2 | 4.1 | 5.7 | 6.2 | 7.6 | 7.7 | 15.9 |
| Intravenous/Subcutaneous Immunoglobulin | 1.2 | 1.2 | 2.4 | 1.2 | 2.4 | 2.4 | 3.5 | 3.5 | 7.1 | 4.7 | 5.9 | 12.9 | 11.8 | 21.2 | 16.5 |
| Vitamin B-12 | 0.8 | 0.9 | 1.6 | 1.6 | 1.5 | 1.4 | 1.9 | 2.6 | 5.1 | 3.6 | 5.8 | 3.6 | 6.1 | 9.2 | 13.3 |
| Manual Lymphatic Drainage | 0.0 | 4.1 | 2.1 | 5.2 | 0.0 | 9.3 | 4.1 | 8.2 | 2.1 | 13.4 | 7.2 | 0.0 | 0.0 | 35.1 | 0.0 |
| Rx Anticoagulants and/or Rx Antiplatelets | 2.0 | 1.0 | 3.4 | 2.0 | 4.4 | 2.5 | 8.8 | 3.9 | 4.9 | 5.9 | 7.8 | 6.4 | 27.5 | 10.3 | 19.6 |
| ADHD Stimulants (Rx) | 0.0 | 0.3 | 3.0 | 1.3 | 1.0 | 2.3 | 1.0 | 1.3 | 4.7 | 4.7 | 4.0 | 4.0 | 4.3 | 9.0 | 12.7 |
| Maraviroc | 1.7 | 0.0 | 0.0 | 0.0 | 3.4 | 0.0 | 1.7 | 0.0 | 1.7 | 0.0 | 1.7 | 3.4 | 15.5 | 6.9 | 17.2 |
| Mestinon | 0.8 | 2.8 | 0.0 | 1.6 | 2.0 | 2.0 | 1.6 | 2.8 | 3.2 | 6.0 | 2.0 | 14.3 | 5.6 | 12.4 | 11.6 |
| Coenzyme Q10 | 0.9 | 0.8 | 1.5 | 1.2 | 1.7 | 1.4 | 1.7 | 2.6 | 4.7 | 4.2 | 5.4 | 4.0 | 6.4 | 9.8 | 14.2 |
| Vagal Nerve Stimulation | 0.0 | 3.0 | 0.0 | 7.6 | 0.0 | 5.3 | 5.3 | 9.8 | 1.5 | 11.4 | 3.8 | 0.0 | 0.0 | 34.8 | 0.0 |
| Palmitoylethanolamide | 1.2 | 1.2 | 1.2 | 0.6 | 2.4 | 1.8 | 0.6 | 1.8 | 4.2 | 3.0 | 5.4 | 7.8 | 3.6 | 12.0 | 14.4 |
| Curcumin | 0.7 | 0.7 | 1.0 | 1.4 | 1.5 | 1.9 | 1.7 | 2.7 | 3.9 | 3.7 | 5.1 | 3.6 | 7.7 | 9.7 | 13.4 |

| Treatment | Craniocervical instability | Adrenal insufficiency or low cortisol | Hypothyroidism | ADD/ADHD | Neuropathy | Other autoimmune disease | Ehlers-Danlos Syndrome or Joint hypermobility | Fibromyalgia | Mast Cell Activation Syndrome | Other dysautonomia | Migraine | Anxiety/Depression | POTS | Orthostatic intolerance |
|---|---|---|---|---|---|---|---|---|---|---|---|---|---|---|
| Fluids/electrolytes | 15.3 | 15.7 | 18.0 | 24.7 | 28.2 | 25.8 | 30.4 | 29.0 | 36.1 | 46.5 | 49.4 | 53.1 | 65.8 | 66.6 |
| Pacing | 7.7 | 8.5 | 20.3 | 13.3 | 0.0 | 21.2 | 21.3 | 34.7 | 20.8 | 32.0 | 50.8 | 55.3 | 44.1 | 59.7 |
| Compression stockings | 17.8 | 16.1 | 18.4 | 26.7 | 31.8 | 28.0 | 35.0 | 30.0 | 40.2 | 50.2 | 50.8 | 51.2 | 74.1 | 72.3 |
| Antihistamines | 16.7 | 16.0 | 17.0 | 26.8 | 30.9 | 26.8 | 31.0 | 26.5 | 45.1 | 45.7 | 48.0 | 52.9 | 57.8 | 57.6 |
| Nattokinase/Lumbrokinase | 16.7 | 17.1 | 15.4 | 24.6 | 33.8 | 25.4 | 28.2 | 15.8 | 40.1 | 46.3 | 38.7 | 45.3 | 57.3 | 50.6 |
| Low-Dose Naltrexone | 19.7 | 19.8 | 23.2 | 22.9 | 27.8 | 29.5 | 33.6 | 32.2 | 39.6 | 47.2 | 51.9 | 51.2 | 60.3 | 67.2 |
| Beta Blockers or Ivabradine | 15.6 | 15.1 | 18.1 | 23.5 | 31.0 | 26.5 | 32.8 | 27.4 | 37.7 | 46.2 | 52.5 | 52.3 | 72.1 | 67.3 |
| Melatonin | 20.0 | 22.8 | 17.5 | 29.6 | 40.5 | 28.7 | 32.2 | 28.0 | 42.9 | 48.6 | 45.9 | 52.0 | 59.5 | 59.9 |
| Other select MCAS treatments | 21.3 | 21.0 | 20.4 | 25.0 | 33.6 | 29.5 | 35.6 | 27.2 | 54.9 | 49.0 | 48.1 | 46.4 | 60.4 | 62.4 |
| Intravenous/Subcutaneous Immunoglobulin | 20.0 | 22.4 | 30.6 | 18.8 | 32.9 | 42.4 | 34.1 | 41.2 | 49.4 | 54.1 | 54.1 | 45.9 | 68.2 | 71.8 |
| Vitamin B-12 | 14.6 | 17.5 | 21.7 | 22.4 | 27.3 | 27.1 | 28.3 | 31.8 | 32.8 | 40.8 | 48.3 | 53.4 | 53.2 | 59.4 |
| Manual Lymphatic Drainage | 11.3 | 13.4 | 20.6 | 16.5 | 0.0 | 16.5 | 17.5 | 50.5 | 28.9 | 32.0 | 58.8 | 55.7 | 43.3 | 60.8 |
| Rx Anticoagulants and/or Rx Antiplatelets | 13.2 | 20.1 | 22.1 | 18.1 | 35.3 | 33.3 | 26.5 | 27.9 | 43.6 | 48.5 | 45.1 | 43.6 | 58.8 | 54.4 |
| ADHD Stimulants (Rx) | 18.3 | 13.7 | 19.3 | 60.0 | 26.3 | 26.0 | 35.7 | 30.0 | 30.0 | 46.7 | 55.3 | 69.7 | 54.7 | 60.0 |
| Maraviroc | 10.3 | 19.0 | 19.0 | 19.0 | 36.2 | 20.7 | 13.8 | 15.5 | 32.8 | 36.2 | 44.8 | 43.1 | 55.2 | 46.6 |
| Mestinon | 18.7 | 19.1 | 17.9 | 21.9 | 33.5 | 27.5 | 40.6 | 32.7 | 47.8 | 49.8 | 53.8 | 41.8 | 78.9 | 86.5 |
| Coenzyme Q10 | 15.8 | 19.3 | 21.4 | 22.6 | 27.4 | 27.7 | 28.7 | 29.7 | 36.4 | 43.3 | 47.6 | 49.8 | 55.2 | 59.8 |
| Vagal Nerve Stimulation | 12.1 | 15.2 | 23.5 | 14.4 | 0.0 | 19.7 | 23.5 | 32.6 | 31.8 | 36.4 | 58.3 | 50.0 | 53.8 | 62.9 |
| Palmitoylethanolamide | 19.8 | 21.6 | 19.2 | 19.8 | 30.5 | 27.5 | 36.5 | 31.7 | 51.5 | 46.1 | 44.3 | 42.5 | 61.1 | 65.9 |
| Curcumin | 16.0 | 18.5 | 19.7 | 22.4 | 31.0 | 30.6 | 27.0 | 34.4 | 36.6 | 45.9 | 46.4 | 52.0 | 51.4 | 57.7 |

Legend scale: 80, 60, 40, 20, 0

For each of the top twenty leading treatments that contributed to improving patients' overall conditions, shown are the percentages of patients who reported on using the treatment and having a certain comorbidity.

**Supplementary Figure 3: Relative Influence of Variables on Treatment Response (NAS)**

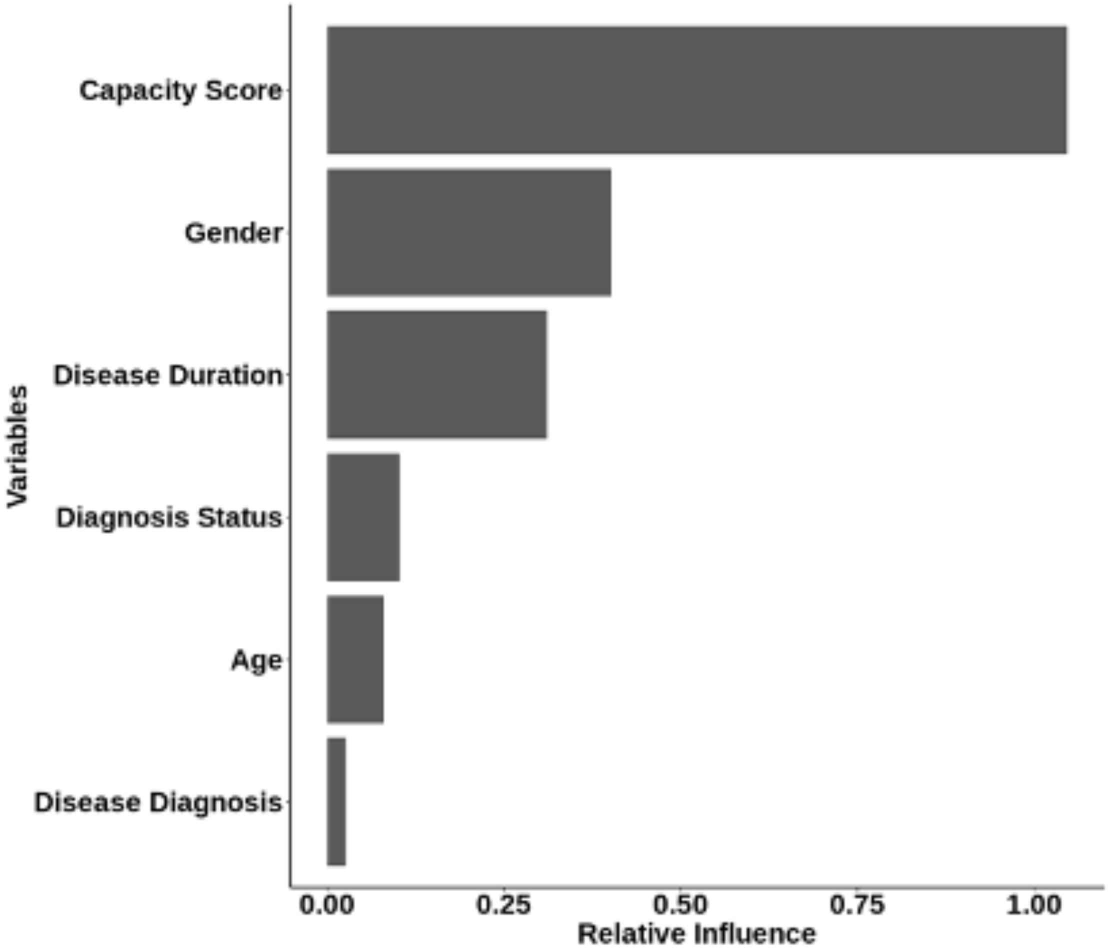


This bar plot illustrates the relative influence of each variable in a Gradient Boosting Machine (GBM) model predicting NAS treatment effectiveness. Variables are listed in order of importance, indicating the strength of their association with the NAS score.

**Supplementary Figure 4. Capacity levels of patients in each cluster**

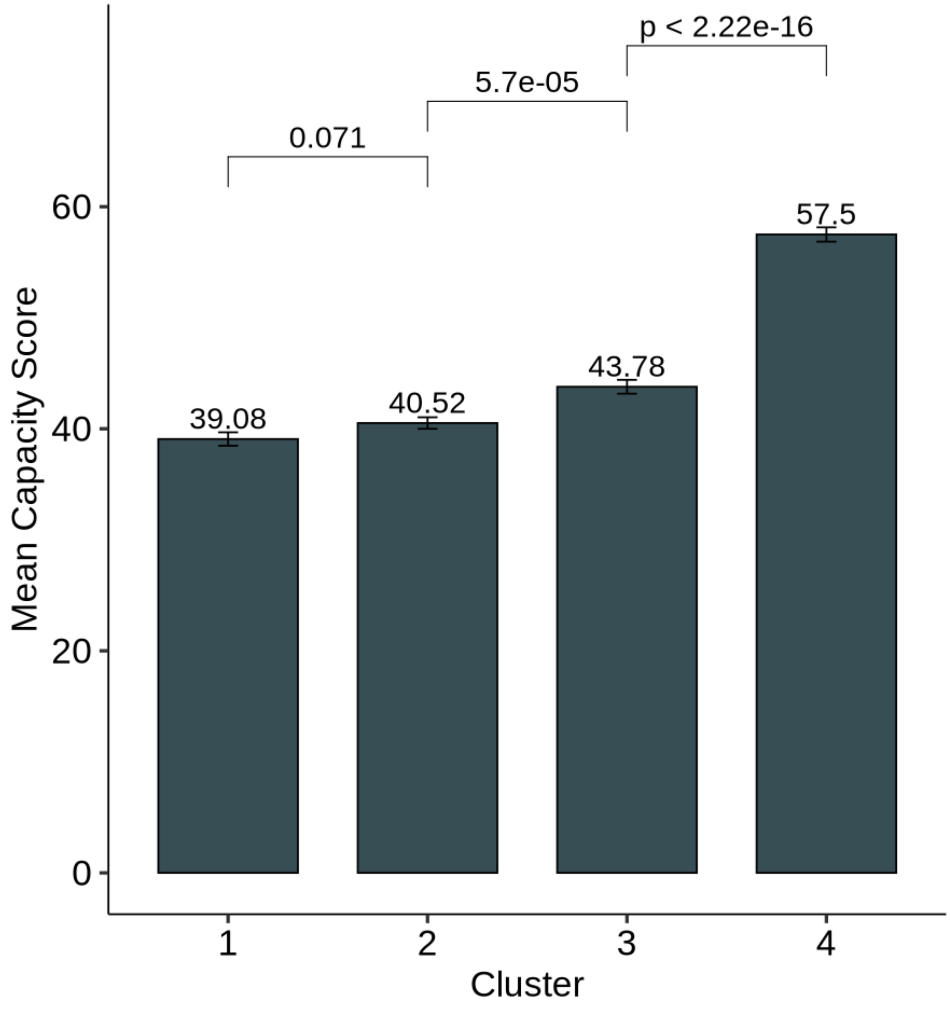


Patient capacity levels decrease from Cluster 4 (least severe symptoms) to Cluster 1 (most severe symptoms).

**Supplementary Figure 5. Patient Subclustering and Symptom Distribution within ME/CFS and Long COVID**

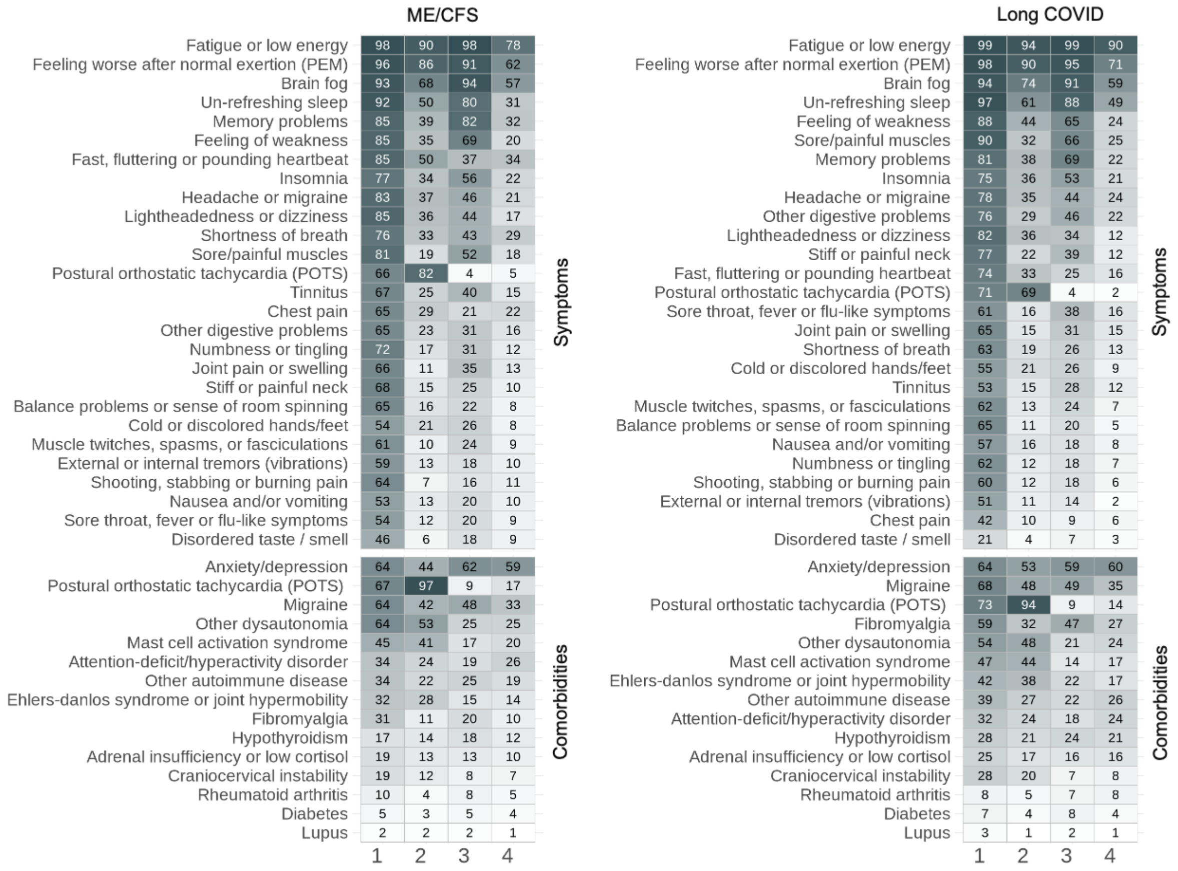

**Supplementary Figure 6 Elbow Method for Determining Optimal Cluster Count in Patient Grouping**

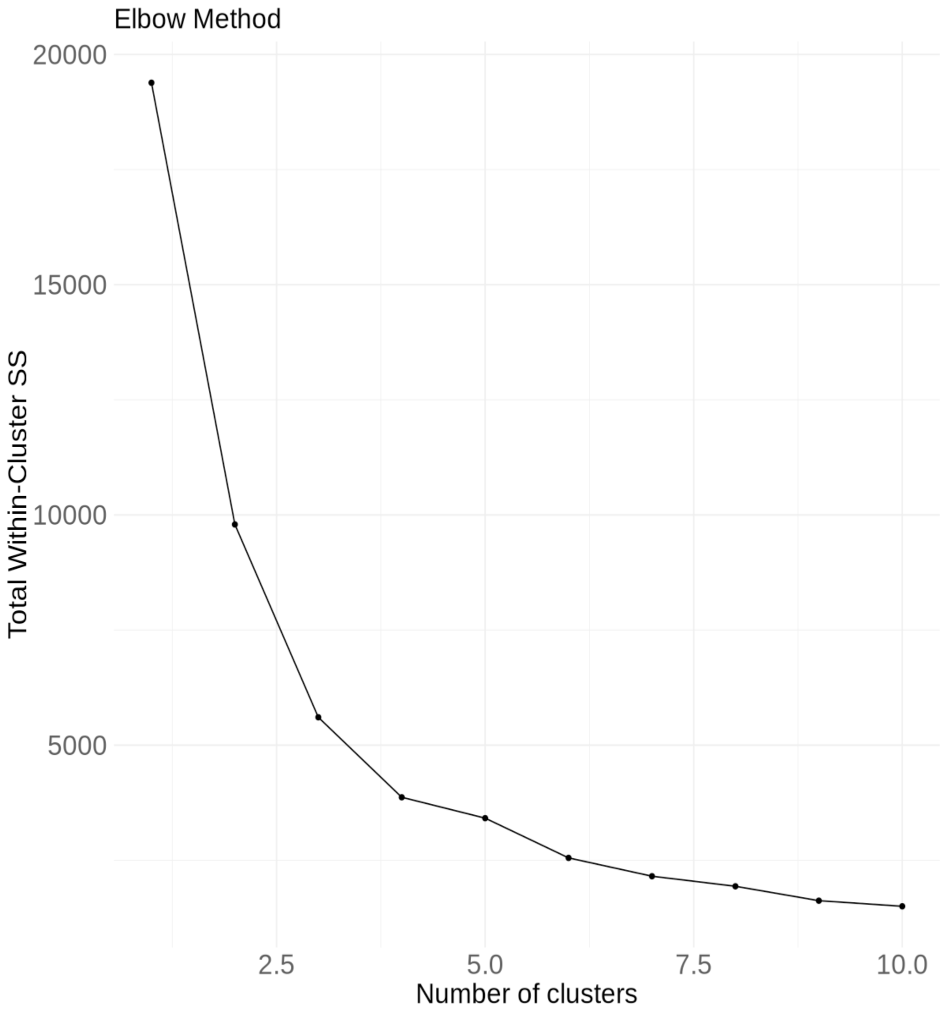
