## Supplementary Document 1 for "Patient-Reported Treatment Outcomes in ME/CFS and Long COVID"

**Question of Patient Capacity Level:**

**How would you describe the CURRENT severity of your ME/CFS?**

*Adapted & condensed from Living with M.E. by Dr Charles Shepherd (ME/CFS Disability Scale, pp. 116-7)*

**FIT & WELL FOR ≥ 1 MONTH** (100% capacity):
No symptoms. Capable of full-time employment.

**GENERALLY WELL** (~90% capacity):
Mild symptoms may sometimes follow activity. Mostly fit & well.

**MILD** (70-90% capacity):
Occasional restriction of capabilities and/or limited ability to perform some tasks requiring exertion. May be able to work full-time.

**MILD TO MODERATE** (50%-70% capacity): 
Variable limited ability to carry out tasks. Able to work part-time and/or perform lighter duties provided adequate rest periods.

**MODERATE** (40-50% capacity): 
Occasionally homebound. Unable to carry out any strenuous duties, but able to perform light tasks for several hours most days.

**MODERATE TO SEVERE** (25-40% capacity): Often homebound. Able to perform light tasks 1-2 hrs some days. May require wheelchair assistance at times.

**SEVERE** (15-25% capacity): 
Bedbound and housebound for much of the time. Considerable difficulties with personal care. Require practical support.

**VERY SEVERE** (<15% capacity):
Bedridden and incapable of living independently. Require a great deal of practical support.

Other (please specify)
